# Mild cognitive impairment is not a valid concept and should be replaced by a more restrictive, biologically validated class, named Mild Cognitive Dysfunctions (MCD): a nomothetic network approach

**DOI:** 10.1101/2021.06.20.21259193

**Authors:** Michael Maes, Sookjaroen Tangwongchai

## Abstract

**Background:** No studies have examined whether interactions between the apolipoprotein E4 (ApoE4) allele and peripheral biomarkers, hypertension, and type 2 diabetes mellitus (T2DM) may impact the neurocognitive, behavioral and social dysfunctions in amnestic mild cognitive impairment (aMCI) and Alzheimer disease (AD).

**Aims:** To clinically define and biologically validate a subgroup of aMCI subjects that take up an intermediate position between controls and AD patients.

**Methods:** In 61 healthy controls, 60 subjects with aMCI, and 60 AD patients we measured the features of aMCI/AD using the Consortium to Establish a Registry for Alzheimer’s Disease (CERAD). A composite BIORISK score was computed using the ApoE4 allele, serum folate, albumin, white blood cells, fasting blood glucose (FBG), atherogenic index of plasma (AIP), T2DM and hypertension.

**Results:** Clustering and nearest neighbour analyses were unable to validate the aMCI subgroup. We constructed two z unit-based composite scores, the first indicating overall burden of cognitive, social, and behavioural deterioration (OBD), and a second reflecting the interactions between ApoE4, all other biomarkers, hypertension and T2DM (BIORISK). We found that 40.2% of the variance in the OBD score was explained by BIORISK, ApoE4, age and education. The OBD index was used to construct three subgroups (normal, medium, and high OBD) with the medium group (n=45) showing mild cognitive dysfunctions (MCD) in memory, language, orientation, and ADL. People with MCD show OBD and BIORISK scores that are significantly different from controls and AD.

**Conclusions:** Petersen’s aMCI criteria cannot be validated and should be replaced by the more restrictive, biologically validated MCD class.

## Introduction

Alzheimer’s disease (AD), the major cause of dementia, is a progressive brain disorder characterized by neuroinflammatory and neurodegenerative processes [1–4]. The early phases of AD are characterized by a gradual decline in neurocognitive functions including impairments in episodic and semantic memory and word fluency [5,6]. In the later stages of AD, patients suffer from deficits in memory, language and naming, orientation, executive functions, perceptual-motor functions, attention, and social skills including communication and judgement [6,7]. At that stage, difficulties to perform activities of daily living (ADL) and neuropsychiatric symptoms, such as behavioral dysregulation, irritability and aggression, inertia, and depressive, vegetative and psychotic symptoms, may be evident [7]. The pathophysiology of AD comprises neuroinflammation with astrogliosis, neurofibrillary tangles, accumulation of amyloid plaques, dystrophic neurites with tau protein, and synaptic, neuronal and neuropil loss [2,3,8,9].

Aging and genetic factors are the most important unmodifiable risk factors for AD and the apolipoprotein E epsilon 4 (ApoE4) allele is the most widely replicated genetic risk factor of AD [7,10]. Around 40% of AD patients carry the ApoE4 allele and risk of AD is increased in E2/E4 (Odds Ratio=2.6), E3/E4 (Odd Ratio=3.2), and especially in E4/E4 (Odds ratio=14.9) carriers [10]. These ApoE genotypes impact the delivery of lipids to cells and the amyloid-β deposits and are associated with increased oxidative stress in the brain and a proinflammatory glial response to inflammatory stimuli [11,12], which play a role in the synaptic dysfunctions, neuroinflammation and neurodegeneration [9,12]. In Thai AD patients, we found that ApoE4 carriers have more impairments in tests of semantic an episodic memory, recall, constructional praxis and praxis recall, naming, clock drawing, ADL functions, and Mini Mental State examination (MMSE), and in communication, language, and judgement [7]. Nevertheless, recent research shows that interactions of the ApoE4 genotype with peripheral biomarkers predict greater impairments in semantic and episodic memory and recall, suggesting that such interactions may play a role in the pathophysiology of AD [13]. For example, interactions between the presence of the E4 allele and fasting blood glucose (FBG) and albumin and cumulative effects of the E4 allele with folic acid, glucose, albumin, and the atherogenic index of plasma (AIP) may increase cognitive deficits in memory and naming and the symptomatic burden in AD [6,13]. Moreover, both hypertension and T2DM may increase risk of AD through immune and oxidative stress associated mechanisms [14–17]. This is important as these peripheral biomarkers, hypertension, and T2DM may increase inflammatory and oxidative pathways thereby aggravating the detrimental effects of the ApoE4 genotypes [6,13]. Nevertheless, no studies have examined whether interactions between those biomarkers, hypertension and T2DM may impact the neurocognitive as well as behavioral and social dysfunctions in AD.

Another unresolved issue is whether similar interaction patterns between the E4 allele and peripheral biomarkers, hypertension, and T2DM may be observed in amnestic mild cognitive impairment (aMCI). Such associations would support that aMCI is a transition stage between normal aging and AD. aMCI is defined as a decline in memory beyond and above that expected by age and the absence of dementia symptoms and dysfunctions in ADL [18]. In the classic classification of aMCI, two subtypes were described, namely single-domain aMCI, with isolated impairments in episodic memory, and multiple-domain aMCI with impairments in episodic memory and one or more other cognitive domains [19]. Individuals with aMCI show an elevated risk to develop AD with a yearly conversion rate from aMCI to AD between 14% and16%, although some aMCI individuals (8%) may return to the normal state [20]. Interestingly, the ApoE4 allele coupled with an interaction between ApoE4 x FBG is associated with memory deficits in people with aMCI [13].

Nevertheless, aMCI as defined by Petersen’s criteria is not a well-modelled entity or nosological class but a heterogenous group of subjects [7]. In fact, there are two main problems with Peterson’s criteria: a) using machine learning techniques such as soft independent modelling of class analogy (SIMCA) performed on neurocognitive test results, we were unable to adequately model the group of people with aMCI using neuropsychological memory tests as modeling variables; and b) using SIMCA, part of aMCI subjects were authenticated as healthy people and another part as AD patients [7]. These results show that using, supervised machine learning techniques, Peterson’s criteria cannot be validated and, consequently, that unsupervised methods should be used to delineate the subgroup of patients who take up an intermediate position between controls and AD patients.

Recently, we developed a new method, namely nomothetic network analysis, to delineate the causal associations between the causome (e.g. ApoE4 allele), the cognitome (the aggregate of all cognitive dysfunctions), and the phenome including the symptomatome (the aggregate of clinical features) of a neuropsychiatric illnesses [21–23]. This conceptual framework may be analyzed using Partial Least Squares (PLS) analysis to define the significant paths between causome, cognitome, and phenome features, followed by unsupervised learning (e.g. cluster analysis), applied to all features of the causal model, to delineate new, more meaningful subgroups.

Hence, the aims of the present study were to a) define the paths from biomarkers (ApoE4, FBG, folate, AIP, albumin, hypertension, and diabetes) and their interactions in association with the cognitome and phenome of aMCI and AD; and b) to define and validate the subgroup of aMCI patients that take up an intermediate position between controls and AD patients with respect to cognitive, behavioral, social and biomarker data.

## Subjects and Methods

### Participants

This is a cross-sectional study which included people with AD and aMCI and normal controls, aged 55-90 years and of both sexes. Patients were recruited at the Dementia Clinic, Outpatient Department, King Chulalongkorn Memorial Hospital, Bangkok, Thailand. Healthy volunteers were recruited from the same catchment area as the patients, namely Patumwan district, Bangkok province. The normal controls were community senior club members, senior Red Cross volunteers, healthy individuals who visited the Health Check Up Clinic, and normal elderly caregivers of the AD patients who visited the Dementia Clinic. We excluded patient and controls with a) abnormal VDRL, HIV, vitamin B12, and thyroid function blood tests; b) other dementia syndromes, including frontotemporal lobe dementia and vascular dementia; c) neurologic disorders, including Parkinson’s disease, stroke, multiple sclerosis, encephalitis, meningitis, and traumatic brain injury; d) major psychiatric disorders including major depressive disorder, bipolar disorder, schizophrenia, substance use disorders, and anxiety disorders; e) (auto)immune disorders, including rheumatoid arthritis, chronic obstructive pulmonary disease, systemic lupus erythematosus, inflammatory bowel disease, chronic kidney disease, cancer, diabetes type 1, and severe heart disease (functional class II or more). Furthermore, controls and patients were excluded when the score on the Thai Geriatric Depression Scale was > 13 in order to exclude people with a recent depression [7]. We conducted magnetic resonance imaging of the brain in the AD patients to rule out vascular dementia and brain tumors.

Finally, all participants were allocated into three study samples, i.e. 61 healthy controls, 60 subjects with aMCI, and 60 AD patients. All participants and guardians of aMCI and AD individuals gave written informed consent prior to participation in this study. The study was conducted according to Thai and international ethics and privacy laws. Approval for the study was obtained from the Institutional Review Board of the Faculty of Medicine,

Chulalongkorn University, Bangkok, Thailand (No 359/56), which is in compliance with the International Guideline for Human Research protection as required by the Declaration of Helsinki, The Belmont Report, CIOMS Guideline and International Conference on Harmonization in Good Clinical Practice (ICH-GCP).

### Clinical measurements

Two senior psychiatrists or neurologists experienced in dementia research assessed patients and controls and completed a semi-structured interview to assess clinical history and diagnostic criteria and they performed neurological and physical examinations, interviewed the close relatives of all participants and measured the Thai version of Clinical Dementia Rating Scale (CDR) [24]. A senior neuropsychologist specialized in dementia assessed neuropsychological test batteries and the Thai Mental Status Examination (TMSE) [25,26]. The neuropsychologist was blinded from the screening data of the physicians and the latter were blinded from the assessment results of the neuropsychologist.

We made the diagnosis of AD using the National Institute of Neurological and Communicative Disorders and Stroke and the Alzheimer’s Disease and Related Disorders Association (NINCDS-ADRDA) diagnostic criteria [27]. Moreover, other inclusion criteria were a TMSE score between 10 and 23, and a CDR score of 1 and 2. aMCI patients were included if they showed subjective memory complaints and when they complied with Peterson’s criteria [18]. Subjective memory complaints were assessed using the question do you feel that your memory had become worse? Objective memory complaints were established with a CDR score of 0.5 and a CDR memory component score of 0.5. aMCI patients were included when the TMSE score was > 23 and when they were not diagnosed with dementia according to the NINCDS-ADRDA) [27]. The healthy controls did not complain of subjective memory and they showed a TMSE score > 23 and CDR = 0. The diagnoses were discussed among two physicians for agreement and in case of disagreement, a third opinion from another psychiatrist or neurologist was requestd.

The same day, a senior neuropsychologist completed the CERAD Neuropsychological Assessment Battery (CERAD-NP) [7,29] in a Thai, validated translation. In this study we used: the Verbal Fluency Test (VFT) to assess semantic memory and cognitive flexibility; the Modified Boston Naming Test (BNT) to assess confrontational word retrieval; the Word List Memory (WLM) to assess episodic memory and learning ability for new verbal information and immediate working memory; WL recall, Delayed, true recall (WLRecall) to probe verbal episodic memory and the ability to recall; the WL Recognition test to assess verbal episodic memory-discriminability or verbal learning recall recognition; and the Constructional Praxis and recall tests to probe visuoconstructive abilities and later task recall. Moreover, we assessed a) C1 or the clinical history items, including memory, language, personality and behavior, orientation for time and place, ADL, social activities, judgment and problem solving, and other cognitive problems; b) C2 or the ADL Blessed Dementia Scale, part a (BDS); C3 or the Behavior Rating Scale for Dementia (BRSD) including depressive features, defective self-regulation, irritability/agitation, vegetative features, inertia/apathy, and psychotic features; C4 or the Short Blessed test (orientation-memory-concentration); and C5 or calculation, clock and expressive language (CCL).

## Biomarker assays

### APOE Genotyping

As described previously [7], we extracted **g**enomic DNA from peripheral blood leukocytes by standard procedures with a DNA Mini Kit (QIAGEN GmbH, Hilden, Germany). Consequently, **“**DNA was amplified by using two primers, 5’-ACAGAATTCGCCCCGGCCTGGTACACAC-3’ and 5’-TAAGCTTGGCACGGCTGAAGGA-3’. Each amplification reaction contained 1 μg of leukocyte DNA, 1 pmol/μl of each primer, 10 % dimethyl sulfoxide, and 0.025 units/pl of *Taq* polymerase in a final volume of 30 μl. Each reaction mixture was heated at 95 °C for 5 min followed by 40 cycles of 95 °C for 60 s, 65 °C for 80 s and 72 °C for 80 s with a final extension at 72 °C for 7 min. The PCR products were treated with ExoSAP-IT (USP Corporation, Cleveland, USA) according to the protocols supplied by the manufacturer, and shipped for direct sequencing to Macrogen Inc. (Seoul, South Korea). In the statistical analyses we used an “ApoE4” group which comprised E4 allele carriers, namely people with the E4/E4 (n=6), E3/E4 (n=32) and E2/E4 (n=5) genotypes [7]. Indeed, one E4 copy (E2/E4 and E3/E4) increases risk for AD and two E4 copies (E4/E4) increased risk considerably [7,10].

### Other biomarkers

Fasting blood was sampled between 8.00 and 8.30 a.m. We used 3 mL clotted blood (serum), which was centrifuged at 1,000 g for 5 minutes, to assay biomarkers at the Central Laboratory, Department of Laboratory Medicine, King Chulalongkorn Memorial Hospital, Bangkok, Thailand. As explained previously [6,13], we used the Architect C8000 (Abbott Laboratories, Abbott Park, Illinois, USA) to measure the biomarkers. Plasma glucose was measured using A Hexakinase/ G-6-PDH technique (inter-assay coefficients of variability (CV) of 2.0%). Based on the lipid profile, we computed the AIP index as a z-unit weighted composite score, i.e. z-transformed triglyceride values (zTG) – z high density lipoprotein chlolesterol (labeled as zAIP) [6]. In addition to albumin, the present study used total number of while blood cells (WBC) as another indicator of immune activation. Folate levels were measured using electrochemiluminescence immunoassay (ECLIA) using the Cobas 6.000 Analyzer (Roche, Germany).

## Statistics

We used analyses of variance (ANOVA) to check differences in scale variables between study groups and analyses of contingence tables (Χ^2^-test) to check associations among categorical variables. We employed multivariate general linear model (GLM) analysis to check the associations between diagnostic classes and clinical and biomarker data while adjusting for age, sex and education. Tests for between-subject effects were used to check the univariate associations between the classes and clinical and biomarker data. Consequently, we computed GLM model-derived estimated marginal means (SE) after adjusting for age, sex and education. We used the protected least significant difference (LSD) to assess pair-wise differences among group means. False discovery rate p-correction was used to correct for multiple comparisons [30]. We used multiple regression analysis to assess the biomarkers that predict latent vector scores while allowing for the effects of age, sex and education. An automated stepwise method was employed with p-to-enter of 0.05 and a p-to-remove of 0.06. Multivariate normality (Cook’s distance and leverage), the R^2^ changes, multicollinearity (using the variance inflation factor and tolerance), and homoscedasticity (tested with the White and modified Breusch-Pagan test) were always checked. Moreover, the regression analysis was performed on 5.000 bootstrap samples and the latter results are shown if the results are not concordant. Tests were 2-tailed and a p-value of 0.05 was used for statistical significance. Two-step cluster analysis was employed to define clusters of patients based on the cognitome and phenome features. Nearest neighbor analysis was employed to classify subjects based on their feature similarities. All statistical analyses were performed using IBM SPSS windows version 25.

Smart Partial Least Squares (SmartPLS)-SEM analysis [31] was used to assess the causal associations between ApoE4, age, sex, education, the cognitome and symptomatome of aMCI and AD. We used a multi-step, multiple mediated PLS path model with ApoE4, age, sex and education as input variables and symptomatome data as output variables, while cognitome data mediated the effects of the input on the output variables. ApoE4, age, sex and education were entered as single indicator variables. Where possible, we entered the cognitome and symptomatome data as latent vectors extracted from the different test and clinical scores. When indicator variables could not be combined in latent vectors, they were entered in the analysis as single indicators. Complete SmartPLS analysis was conducted when the outer and inner models complied with specific pre-specified quality criteria, namely: a) the overall model fit SRMR is < 0.08; b) the vector loadings are all > 0.666 at p < 0.001; c) the outer model latent vectors show a good construct validity, namely composite reliability > 0.7 and average variance extracted (AVE) > 0.5; and d) Confirmatory Tetrad Analysis (CFA) shows that the latent vector models constructed as reflective models are not mis-specified. Complete PLS-SEM analysis performed on 5.000 bootstrap samples was used to compute outer model loadings and path coefficients with p values and specific indirect and total effects. The predictive power of the model was assessed using blindfolding and PLSpredict with 10-fold cross-validation.

## Results

### Results of PLS analysis

**Figure 1** shows the final PLS model. The symptomatome was structured in 4 different vectors, namely ADL+OR (BDSa, SBT score and C1 orientation), BEHAVIOR (the C3 BRSD items comprising depressive features, irritability/agitation, vegetative features, inertia/apathy, and defective self-regulation), MEM+LANG (comprising the C1 clinical history items memory and language), and SOCIAL (including the C1 clinical history items social activities, judgment and problem solving, and other cognitive problems). We also constructed two neurocognitive test latent vectors, namely a) a CERAD latent vector comprising the scores on the BNT, VFT, WLM, WLRecall, WLRecognition, Constructional Praxis and Constructional Praxis Recall; and b) a latent vector comprising calculation, clock drawing, and expressive language (CCL latent vector). In the mediation model, CCL and CERAD mediated the effects of age, ApoE4 and education (entered as single indicator input variables) on the 4 symptomatome latent vectors. Finally, the 4 symptomatome latent vectors were used as predictors of the diagnosis (entered as 0, 1 and 2 for controls, aMCI and AD, respectively). The model displayed in Figure 1 shows an adequate model fit with SRMR=0.057. The construct reliabilities of all 6 latent vectors are adequate with all AVE values > 0.542 and all composite reliabilities > 0.755. Moreover, all loadings on the 7 latent vectors were all > 0.66 at p<0.0001 and the vectors were not mis-specified as reflective models. Blindfolding showed that the construct cross-validated redundancies of all constructs were adequate. All Q^2^Predict scores of the indicators were positive indicating that they outperform the most naïve benchmark.

**Figure 1.**
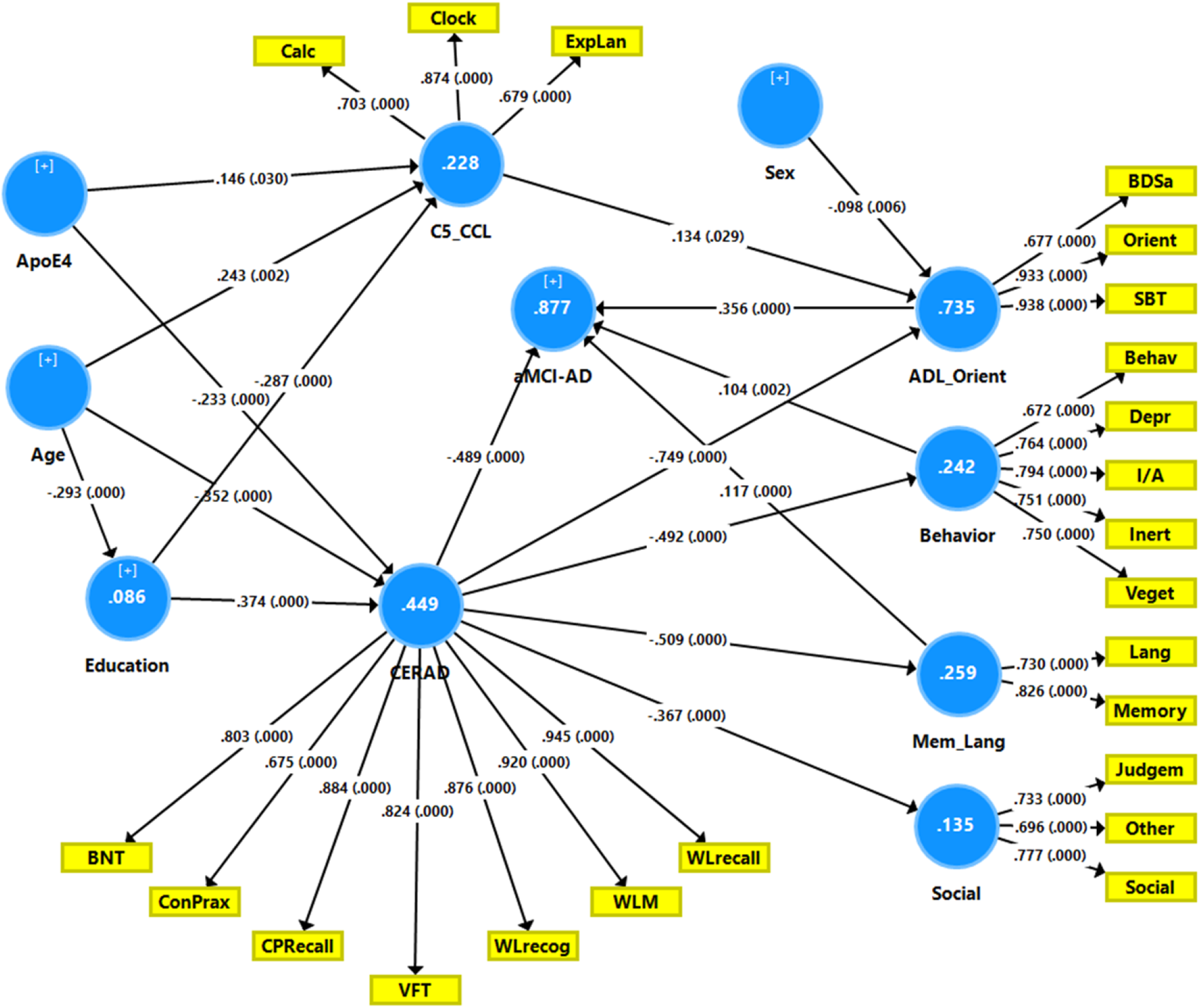
Results of complete Partial Least Squares analysis performed on 5.000 bootstrap samples. Shown are path coefficients and loadings (with p values). White figures within the circles indicate percentage of variance explained. The ApoE4 allele, age, sex and education are input variables and symptomatome data are output variables with cognitome data mediating the effects of the input on the output variables. CCL: calculation (calc), clock, expressive language (ExpLan). CERAD: a latent vector extracted from various CERAD scores including BNT (Boston Naming Test), ConPrax (Constructional Praxis) and recall (CPRecall), VFT: verbal fluency test; WLRecog: Word List Recognition; WLM: Word List Memory; and WLRecall. ADL_Orient: activities of daily living and orientation, comprising the Blessing Dementia Scale part a (BDSa), SBT: Short Blessing Test, and C1 orientation. Behavior: Behavior Rating Scale for Dementia subdomains, including depressive features (depr), defective self-regulation (beh), irritability/agitation (I/A), vegetative features (veget), inertia/apathy (inert). Mem_Lang: C1 clinical history items, including memory and language (Lang). Social: C1 clinical items, including social activities, judgment and problem solving, and other cognitive problems.

We found that 87.7% of the variance in the diagnostic variable was explained by (in ascending order of importance) CERAD, ADL+OR, MEM+LANG, and BEHAVIOR, while CCL and SOCIAL did not have a significant impact. PLS showed that 44.9% of the variance in CERAD could be explained by ApoE4, age, and education, and that the same indicators explained 22.8% of the variance in CCL. Moreover, the effects of CERAD and CCL on the diagnostic spectrum was partially mediated by ADL+OR, BEHAVIOR, and SOCIAL. The specific indirect effects showed that most pathways from the input to the output variables were significant. For example, the effects of ApoE4, age and education on the diagnosis were mediated by CERAD, or CERAD → ADL+OR, or CERAD → BEHAVIOR, CERAD → MEM+LANG, but not CCL → ADL_OR. The significant specific indirect effects of sex on the diagnosis were mediated by ADL+OR. In addition, all possible total effects were significant with total effects of ApoE4 on CERAD, CCL, ADL+OR, BEHAVIOR, SOCIAL, and MEM+LAN.

### Results of nearest neighbor analyses

Two different nearest neighbor analyses were performed to classify subjects as controls, aMCI or AD patients. The first was conducted using memory scores only, namely WLM, WLRecall and C1 memory scores (3k, Euclidian distance, training sample of 70% and a holdout sample of 30%). The classification table showed that many aMCI cases were misclassified as controls in the training (35.7%) and holdout sample (45.4%) yielding a total accuracy of only 68.9% in the holdout sample. The second analysis was conducted using all cognitome and phenome latent vectors extracted by PLS scores (3k, Euclidian distance, training sample of 70% and a holdout sample of 30%) and this analysis showed 45.0% misclassifications in both the training and holdout samples with many aMCI subjects being allocated to the normal control class.

### Construction of an overall burden of disease (OBD) score and associated subgroups

We computed the latent variable scores of all cognitome, and phenome data obtained by PLS and computed an overall composite score indicating overall burden of disease (OBD) computed as: z score of (z CCL + z ADL+OR + z BEHAVIOR + z MEM+LANG + z SOCIAL – z CERAD). Consequently, using a visual binning method (based on inspection of the apparent modes and local minima of the frequency histogram and the results of the two-step cluster analysis described below), we divided the study group into three non-overlapping samples, namely normal, medium and high OBD (cutoff points were −0.53 and 0.4, respectively). Two-step cluster analysis performed on CCL, CERAD, ADL+OR, MEM_LANG, BEHAVIOR, SOCIAL, and OBD scores in the normal + medium OBD groups retrieved 2 clusters (based on Akaike’s Information criterion) with an adequate silhouette measure of cohesion and separation of 0.57. This cluster solution separated the medium OBD from the normal OBD class. **Table 1** shows the association between this new OBD and cluster analysis classification and the classification into HC, aMCI and AD. There was a highly significant association between both classification systems (χ^2^=206.97, df=4, p<0.001), whereby group 1 comprised most controls (except 2) + 21 aMCI subjects (this group was labeled “normal OBD group”), group 2 consisted of 36 aMCI + 2 healthy controls + 7 AD subjects (labeled:” medium OBD group”), and group 3 comprised 53 AD + 3 aMCI subjects (labeled “high OBD group”).

**Table 1.** Associations between the classes derived from cluster analysis and the diagnosis of amnestic mild cognitive impairment (aMCI) and Alzheimer’s disease.

| **Classes** | **Normal OBD <sup>A</sup>** | **Medium OBD <sup>B</sup>** | **High OBD <sup>C</sup>** | **Total** |
| --- | --- | --- | --- | --- |
| HC | 59 | 2 | 0 | 61 |
| aMCI | 21 | 36 | 3 | 60 |
| AD | 0 | 7 | 53 | 60 |
| Total | 80 | 45 | 56 | 181 |

OBD: overall burden of disease classes (computed based on the results of cognitive, behavioral and social rating scores).
HC: healthy controls; aMCI: amnesic mild cognitive impairment, AD: Alzheimer's disease.

### Clinical and biological features of the OBD classes

**Table 2** shows the features of the 3 OBD groups formed. Patients with high OBD are somewhat older that the other groups and years of education was lower. There were no differences in the sex ratio or cardio-vascular disease frequency among the three study groups. The frequency of hypertension increased from controls → medium OBD → high OBD and that of type 2 diabetes mellitus (T2DM) was higher in both OBD groups than in controls. The frequency of ApoE4 was significantly higher in the high OBD group than in the normal OBD group.

**Table 2.** Socio-demographic, clinical and biomarker data of the participants binned into three groups based on the overall burden of disease (OBD) score and cluster analysis.

| Variables | Group 1 or normal OBD (n=80) <sup>A</sup> | Group 2 or medium OBD (n=45) <sup>B</sup> | Group 3 or high OBD (n=56) <sup>C</sup> | F/FEPT/$\chi^2$ | df | p |
| --- | --- | --- | --- | --- | --- | --- |
| Age (years) | 69.5 ± 6.1 <sup>B,C</sup> | 75.2 ± 6.9 <sup>A,C</sup> | 79.0 ± 6.7 <sup>A,B</sup> | 36.05 | 2/178 | <0.001 |
| Sex | 18/62 | 11/34 | 17/39 | 1.10 | 2 | 0.576 |
| Education (years) | 12.4 ± 5.0 <sup>B,C</sup> | 9.5 ± 5.9 <sup>A,C</sup> | 6.2 ± 5.0 <sup>A,B</sup> | 23.01 | 2/178 | <0.001 |
| CVD (N/Y) | 75/5 | 41/4 | 48/8 | 2.52 | 2 | 0.284 |
| Hypertension (N/Y) | 61/19 <sup>B,C</sup> | 25/20 <sup>A,C</sup> | 15/41 <sup>A,B</sup> | 32.68 | 2 | <0.001 |
| T2DM (N/Y) | 72/8 <sup>B,C</sup> | 33/12 <sup>A</sup> | 42/14 <sup>A</sup> | 7.30 | 2 | 0.026 |
| Allele E4 (N/Y) | 71/9 <sup>C</sup> | 34/11 | 33/23 <sup>A</sup> | 16.19 | 2 | <0.001 |
| CCL (z scores) * | -0.531 ± 0.091 <sup>B,C</sup> | -0.248 ± 0.107 <sup>A,C</sup> | 0.959 ± 0.110 <sup>A,B</sup> | 49.27 | 2/175 | <0.001 |
| CERAD (z scores) * | 0.751 ± 0.059 <sup>B,C</sup> | 0.054 ± 0.069 <sup>A,C</sup> | -1.117 ± 0.071 <sup>A,B</sup> | 167.62 | 2/175 | <0.001 |
| ADL+OR (z scores) * | -0.751 ± 0.059 <sup>B,C</sup> | -0.261 ± 0.069 <sup>A,C</sup> | 1.282 ± 0.071 <sup>A,B</sup> | 210.39 | 2/175 | <0.001 |
| Behavior (z scores) * | -0.460 ± 0.107 <sup>C</sup> | -0.221 ± 0.125 <sup>C</sup> | 0.834 ± 0.129 <sup>A,B</sup> | 27.18 | 2/175 | <0.001 |
| Memory+Language (z scores) * | -0.703 ± 0.099 <sup>B,C</sup> | 0.207 ± 0.117 <sup>A,C</sup> | 0.838 ± 0.120 <sup>A,B</sup> | 40.50 | 2/175 | <0.001 |
| Social (z scores) * | -0.461 ± 0.109 <sup>C</sup> | -0.299 ± 0.128 <sup>C</sup> | 0.900 ± 0.132 <sup>A,B</sup> | 30.62 | 2/175 | <0.001 |
| OBD index (z scores) * | -0.819 ± 0.048 <sup>C</sup> | -0.196 ± 0.056 <sup>C</sup> | 1.327 ± 0.057 <sup>A,B</sup> | 345.26 | 2/175 | <0.001 |
| FBG (z scores) | -0.247 ± 0.812 <sup>C</sup> | 0.062 ± 0.927 | 0.304 ± 0.1204 <sup>A</sup> | 5.39 | 2/178 | 0.005 |
| Albumin (z scores) | 0.201 ± 0.923 <sup>C</sup> | 0.084 ± 0.896 <sup>C</sup> | -0.361 ± 1.098 <sup>A,B</sup> | 5.75 | 2/178 | 0.004 |
| White blood cells (z scores) | -0.240 ± 0.856 <sup>C</sup> | -0.060 ± 1.077 <sup>C</sup> | 0.391 ± 1.025 <sup>A,B</sup> | 7.11 | 2/178 | 0.001 |
| Folate (z scores) | 0.242 ± 0.901 <sup>C</sup> | 0.111 ± 0.996 <sup>C</sup> | -0.436 ± 1.012 <sup>A,B</sup> | 8.61 | 2/178 | <0.001 |
| zAIP (z scores) | -0.184 ± 0.999 <sup>C</sup> | -0.092 ± 1.020 <sup>C</sup> | 0.337 ± 0.914 <sup>A,B</sup> | 4.94 | 2/178 | 0.008 |
| zBiomarkers (z scores) | -0.453 ± 0.730 <sup>B,C</sup> | -0.089 ± 0.885 <sup>A,C</sup> | 0.720 ± 1.023 <sup>A,B</sup> | 30.38 | 2/178 | <0.001 |
| zBIORISK | -0.498 ± 0.708 <sup>B,C</sup> | -0.018 ± 0.889 <sup>A,C</sup> | 0.725 ± 1.011 <sup>A,B</sup> | 33.56 | 2/178 | <0.001 |
| zBIORISK+OBD | -0.761 ± 0.432 <sup>B,C</sup> | -0.123 ± 0.522 <sup>A,C</sup> | 1.191 ± 0.694 <sup>A,B</sup> | 212.06 | 2/178 | <0.001 |

All variables are shown as mean (SD) except \*: marginal estimated mean (SE) after covarying for age, sex and education.
<sup>A,B,C</sup> pairwise comparisons among treatment means.
CVD: cardio-vascular disease; T2DM: type 2 diabetes mellitus; CCL: calculation, clock, expressive language; CERAD: a latent vector extracted from various CERAD scores including memory, naming and praxis; ADL+OR: activities of daily living and orientation; OBD: overall burden of cognitive, behavioral and social deterioration; FBG: fasting blood glucose; zAIP: atherogenic index of plasma.
zBiomarkers: z unit weighted composite score computed as z ApoE4 + z white blood cells + z FBG + z AIP – z Albumin – z Folate.
zBIORISK: computed as zBiomarkers + z hypertension + z T2DM.

In order to assess the associations between these groups and the clinical and neuropsychological scores we used multivariate GLM analysis and adjusted for age, sex and education. There was a highly significant effect of the diagnostic classes (partial eta squared=0.629; F=41.01, df=14/340, p<0.001) and a very modest effect of education (partial eta squared=0.082; F=2.15, df=7/169, p=0.041), while sex and years of education did not have significant effects. Tests of parameter estimates shows that education was only associated with the CERAD score (inversely, p=0.005). The CCL, CERAD, ADL+OR and MEM+LANG latent variable scores increased from normal OBD → medium OBD → high OBD. The BEHAVIOR and SOCIAL scores were significantly increased in the high OBD group as compared with the other two groups. These differences remained significant after FDR p-correction. Table 2 also shows the measurement of blood biomarkers in the three study groups. FBG was significantly higher in the high OBD group than in the normal and medium OBD group. Albumin and folate were significantly lower in the high OBD group than in the normal and medium OBD group, while WBC number and zAIP were significantly increased in the high OBD group as compared with the other two groups. FDR p-correction did not change these results.

We have also computed a z unit weighted composite score that comprises all biomarkers (zBiomarkers) as a z ApoE4 + z WBC + z FBG + z AIP – z Alb – zFolate (labeled as: zBiomarkers). In addition, we computed another z unit-based composite score as: zBiomarkers + z hypertension + z T2DM (labeled as zBIORISK). Table 2 shows that both indices were significantly different between the three groups and increased from the normal OBD → medium OBD → high OBD group. The distance from the medium OBD to the normal OBD group was 0.480 SD and between the medium and high OBD the distance was 0.743 SDs. Finally, we calculated a composite score as zBIORISK + zOBD score. The distance from the medium OBD to the normal OBD group was 0.641 SDs and between the medium and high OBD the distance was 1.314 SDs indicating that the medium OBD group takes up an intermediate position between the normal and high OBD groups, but is much closer to the normal OBD group than to the high OBD group.

### Associations between biomarkers and the OBD indices

**Table 3** shows the intercorrelation matrix between the zBiomarker and zBIORISK factors and the clinical and neuropsychological scores. We found that these risk indices were significantly associated with CCL, CERAD, ADL+OR, BEHAVIOR, MEM+LANG, and SOCIAL scores. **Table 4** shows the results of multiple regression analysis with the OBD score as dependent variable and the biomarker indices as explanatory variables while allowing for the effects of age, sex and education. Regression #1 shows that 40.2% of the variance in OBD was explained by the regression on zBIORISK, age, ApoE4 (all positively) and education (inversely). **Figure 2** shows the partial regression of the OBD score on the zBIORISK score. We have also examined the effects of the biomarker score on the OBD score in restricted samples, namely normal + medium OBD and medium + high OBD groups. Regression #2 shows that in the restricted study sample of normal + medium OBD subjects, 19.5% of the variance in the OBD score could be explained by zBIORISK and age (both positively) and education (inversely). Regression #3 shows that in the medium + high OBD group, 17.0% of the variance in the OBD score could be explained by zBiomarkers (positively) and education (inversely).

**Table 3.**
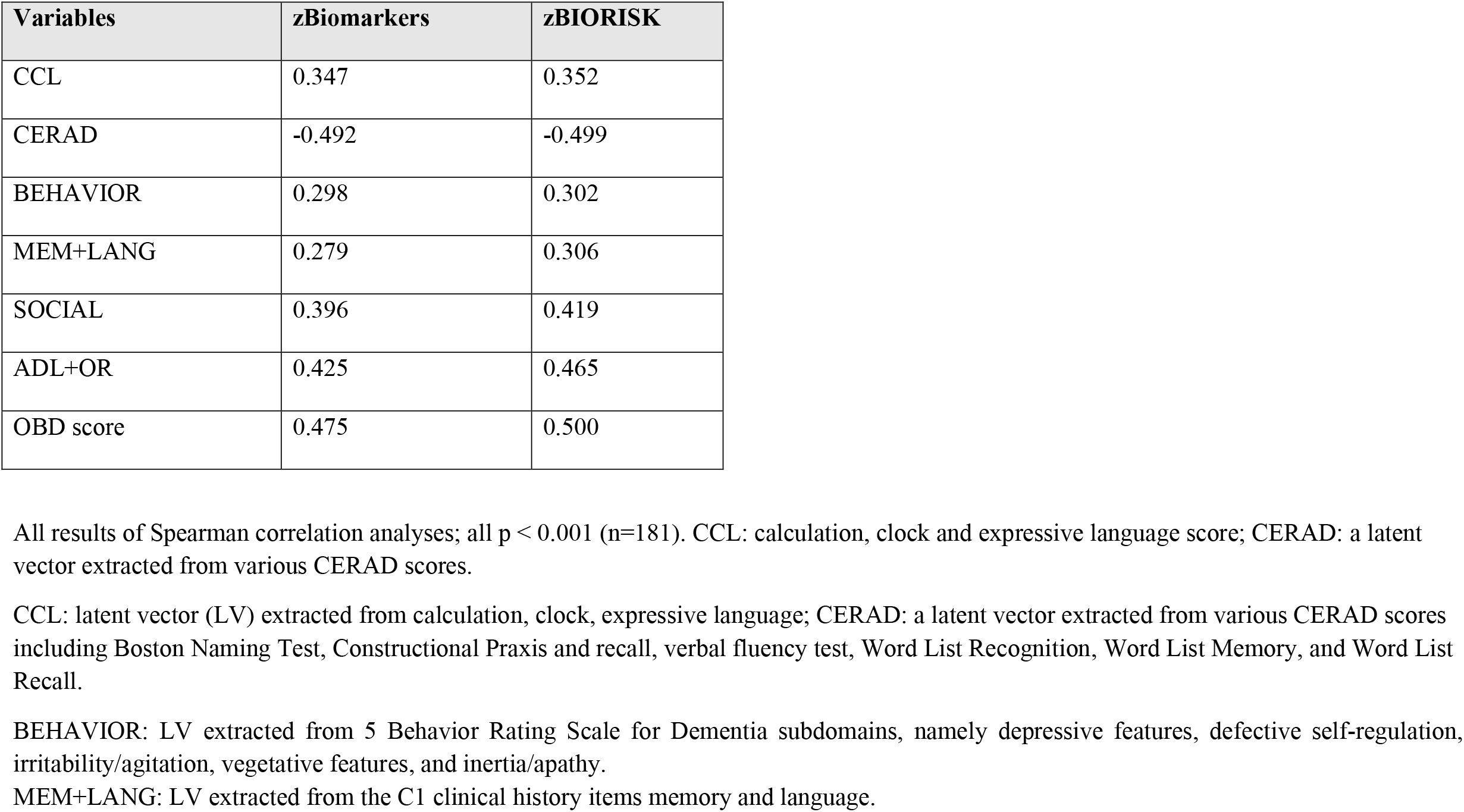
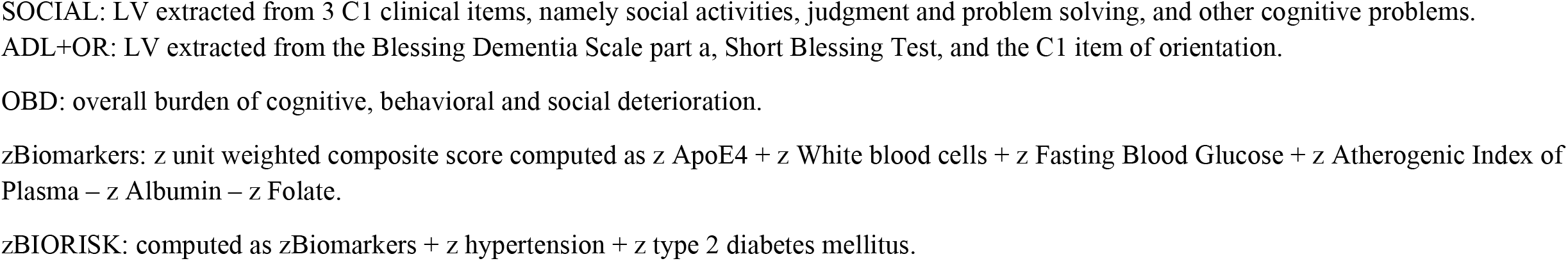
Intercorrelation matrix between biomarker and clinical and cognitive scores

| **Variables** | **zBiomarkers** | **zBIORISK** |
| --- | --- | --- |
| CCL | 0.347 | 0.352 |
| CERAD | -0.492 | -0.499 |
| BEHAVIOR | 0.298 | 0.302 |
| MEM+LANG | 0.279 | 0.306 |
| SOCIAL | 0.396 | 0.419 |
| ADL+OR | 0.425 | 0.465 |
| OBD score | 0.475 | 0.500 |

All results of Spearman correlation analyses; all $p < 0.001$ ( $n=181$ ). CCL: calculation, clock and expressive language score; CERAD: a latent vector extracted from various CERAD scores.
CCL: latent vector (LV) extracted from calculation, clock, expressive language; CERAD: a latent vector extracted from various CERAD scores including Boston Naming Test, Constructional Praxis and recall, verbal fluency test, Word List Recognition, Word List Memory, and Word List Recall.
BEHAVIOR: LV extracted from 5 Behavior Rating Scale for Dementia subdomains, namely depressive features, defective self-regulation, irritability/agitation, vegetative features, and inertia/apathy.
MEM+LANG: LV extracted from the C1 clinical history items memory and language.
SOCIAL: LV extracted from 3 C1 clinical items, namely social activities, judgment and problem solving, and other cognitive problems.
ADL+OR: LV extracted from the Blessing Dementia Scale part a, Short Blessing Test, and the C1 item of orientation.
zBiomarkers: z unit weighted composite score computed as $z \text{ ApoE4} + z \text{ White blood cells} + z \text{ Fasting Blood Glucose} + z \text{ Atherogenic Index of Plasma} - z \text{ Albumin} - z \text{ Folate}$ .
zBIORISK: computed as $z\text{Biomarkers} + z \text{ hypertension} + z \text{ type 2 diabetes mellitus}$ .

**Table 4.** Results of multiple regression analyses with the overall burden of cognitive, behavioral and social deterioration (OBD) score as dependent variable and biomarker scores as explanatory variables.

| Dependent variables | Explanatory Variables | $\beta$ | t | p | F model | df | p | R<sup>2</sup> |
| --- | --- | --- | --- | --- | --- | --- | --- | --- |
| #1. OBD in all | **Model** |  |  |  | 29.54 | 4/176 | <0.001 | 0.402 |
|  | zBIORISK | 0.261 | 3.72 | <0.001 |  |  |  |  |
|  | Education | -0.256 | -4.17 | <0.001 |  |  |  |  |
|  | Age | 0.260 | 3.87 | <0.001 |  |  |  |  |
|  | ApoE4 | 0.1349 | 2.11 | 0.036 |  |  |  |  |
| #2. OBD in normal and medium OBD groups | **Model** |  |  |  | 9.43 | 3/117 | <0.001 | 0.195 |
|  | zBIORISK | 0.193 | 2.05 | 0.043 |  |  |  |  |
|  | Education | -0.227 | -2.70 | 0.008 |  |  |  |  |
|  | Age | 0.225 | 2.41 | 0.017 |  |  |  |  |
| #3. OBD in medium and high OBD groups | zBiomarkers | 0.340 | 3.67 | <0.001 | 10.01 | 2/98 | <0.001 | 0.170 |
|  | Education | -0.188 | -2.19 | 0.031 |  |  |  |  |

zBiomarkers: z unit weighted composite score computed as z ApoE4 + z White blood cells + z Fasting Blood Glucose + z Atherogenic Index of Plasma – z Albumin – z Folate.
zBIORISK: computed as zBiomarkers + z hypertension + z type 2 diabetes mellitus.
ApoE4: any of the E2/E4, E3/E4 or E4/E4 genotypes.

**Figure 2.**
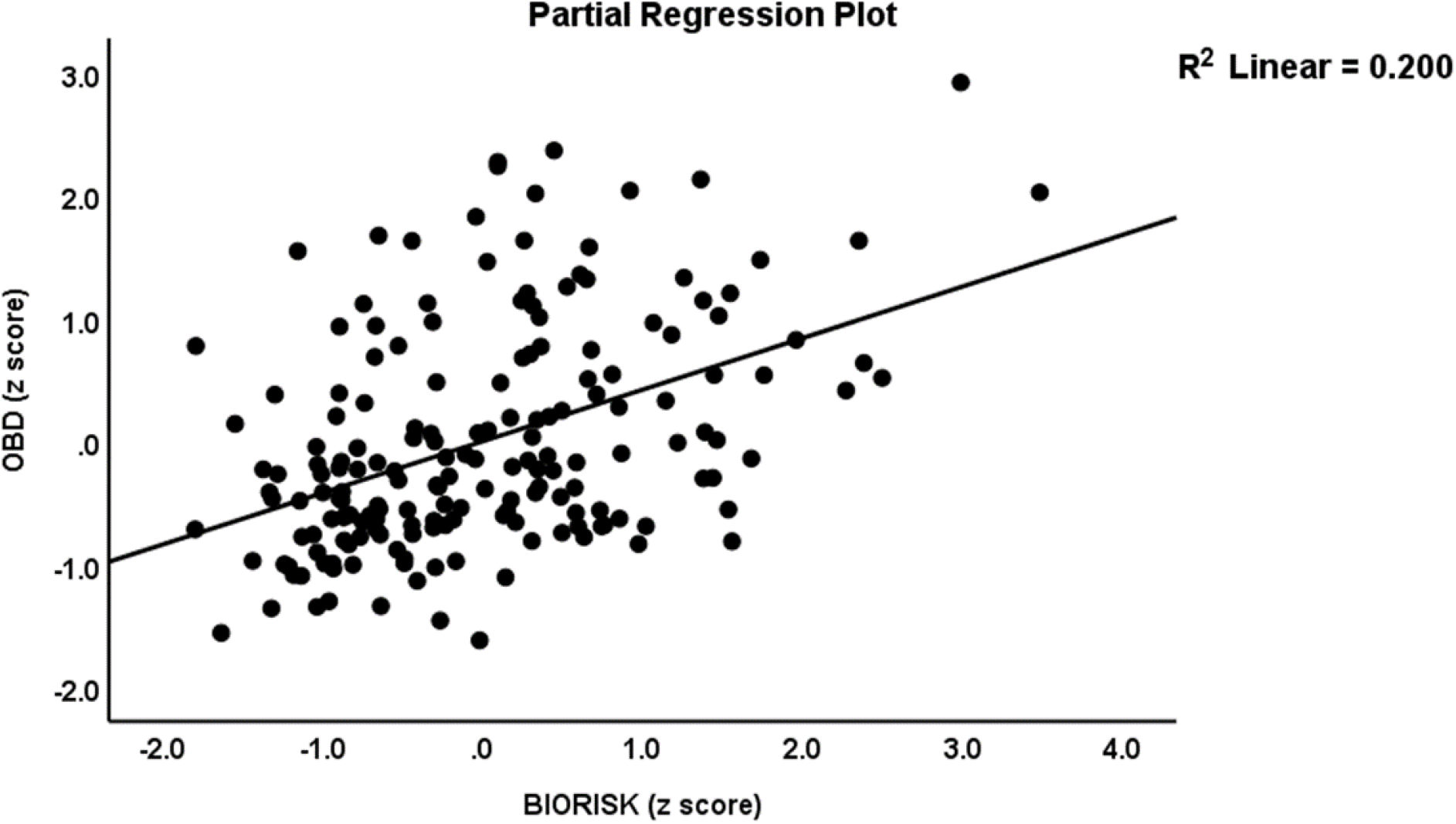
Partial regression of the overall burden of cognitive, behavioral and social deterioration (OBD) on the BIORISK index. The OBD index was computed using cognitive, behavioural, and social symptoms of the CERAD. The BIORISK score was computed using the ApoE genotypes, serum folate, albumin, white blood cells, fasting blood glucose, atherogenic index of plasma, type 2 diabetes mellitus, and hypertension.

## Discussion

### aMCI cannot be validated as a class

The first major finding of this study is that nearest neighbor (supervised learning) and two-step clustering (unsupervised learning) analysis were not able to confirm the existence of aMCI even when using memory scores, which defined aMCI. These results corroborate those of our previous report showing that using SIMCA, another supervised learning technique [7], the aMCI subgroup cannot even be modeled using neuropsychological memory tests, which are intended to describe this subgroup [18]. Moreover, using SIMCA, we found that many participants with aMCI were authenticated as controls or as AD patients. Thus, both unsupervised and supervised techniques show that aMCI according to Peterson’s criteria is a heterogenous group and, consequently, does not exist as a distinct class. In fact, this is further corroborated by the low diagnostic performance of different neuropsychological tests when discriminating aMCI subjects from controls. These figures show an accuracy of around 70-80% as reviewed in [32], where in fact a boostrapped accuracy of > 95% would be needed to obtain a good separation. Using the most adequate machine learning techniques to classify subjects (including support vector machine or neural network analysis) did not improve these figures considerably [32,33].

### Mild Cognitive Dysfunctions (MCD) as an intermediate class

The second major finding of this study is that we were able to compute a new overall burden of cognitive, social and behavioural deterioration (OBD) score, which is useful as a severity score and to delineate a reliable subgroup located between controls and AD patients. This OBD index combines the different cognitome (CCL and CERAD) and phenome (ADL+OR; BEHAVIOR, MEM+LANG, SOCIAL) domains of the dementia spectrum into one integrated index of overall burden of disease and, therefore, ranks subjects along a continuum from a normal condition to severe dementia. Using a visual binning method performed on the OBD scores, we divided the study sample into three classes, namely subjects with normal, high and medium OBD scores and validated the medium OBD class using two-step cluster analysis. This medium OBD group is less inclusive than aMCI and is characterized by impairments in CERAD, CCL, ADL+OR and MEM+LANG and, therefore, we propose to name this group Mild Cognitive Dysfunctions (MCD) including in memory, ADL, language and orientation. Peterson’s criteria [18] on the other hand, stress the absence of ADL dysfunctions in aMCI. As such, the features of MCD differ from those of single-domain aMCI and of multiple-domain aMCI, which was thought to be characterized by deficits in episodic memory and one or more other cognitive domains [19]. Moreover, the MCD criteria do not correspond with those of mild behavioral impairment (MBI) which is characterized by persistent behavioral symptoms in late life and is thought to constitute a risk for neurodegenerative disease [34,35]. Indeed, in our study the MCD group did not display any behavioral or social dysfunctions.

### Biomarkers in MCD

The third major finding of this study is that ApoE4 significantly predicted all cognitome (CCL and CERAD) and phenome (ADL+OR; BEHAVIOR, MEM+LANG, SOCIAL) domains of the dementia spectrum, and that the impact of the ApoE4 allele could be improved by constructing a new composite score reflecting the interactions between the ApoE4 allele, folate, FBG, albumin, WBC, and AIP, and comorbid illness including hypertension and T2DM. This biomarker score externally validated the continuous OBD score and the MCD class: a) people with MCD show higher biomarker scores than controls, and b) the transitions of controls to MCD and from the latter to AD are both associated with increasing biomarker scores. Phrased differently, an increased impact of interactions between factors which confer risk towards increased glucotoxicity (ApoE4 x FBG x T2DM), atherogenicity (ApoE4 x AIP x hypertension), inflammatory responses (ApoE4 x albumin x WBC), and oxidative stress (ApoE4 x folate x albumin) underpin both MCD and AD. Since the same biomarker score is associated with AD as well as with MCD, we may conclude that people with MCD show probably an increased risk to develop AD. It is known that individuals with aMCI display an increased risk to develop AD with a conversion rate from aMCI to AD of 14%-16.5% [20]. Future research should examine how many MCD subjects show the expected conversion rate to develop AD.

## Limitations

The results of this study should be interpreted with regard to the limitations. It would have been more interesting if we had assayed brain imaging biomarkers including the connectome and neuro-immune biomarkers which are known to play a role in AD and in cognitive deterioration including levels of neurotoxic cytokines and chemokines and oxidative stress biomarkers as well as more specific antioxidants (Rottkamp et al., 2000; Huang et al., 2016; Morris et al., 2019).

## Conclusions

Both supervised and unsupervised learning techniques show that aMCI is a heterogenous class and not a viable entity. In this study, we constructed two z unit-based composite scores: a first reflecting overall burden of cognitive, social and behavioural deterioration (OBD) and a second score reflecting the interactions between ApoE4 and other biomarker risk factors, hypertension and T2DM. The OBS score may be used to assess severity of OBD and AD and to classify MCD subjects. The latter show increased biomarker scores which significantly differ from controls and AD patients and, therefore, the MCD class may be at increased risk to develop dementia.

## Data Availability

The dataset generated during and/or analyzed during the current study will be available from the corresponding author upon reasonable request and once the dataset has been fully exploited by the authors.

## Authorships

All authors contributed significantly to the paper and approved the final version.

## References

1. Fratiglioni L, De Ronchi D, Agüero-Torres H. Worldwide prevalence and incidence of dementia. Drugs Aging 1999;15(5):365–75. doi: 10.2165/00002512-199915050-00004. PMID: 10600044.

2. Edler MK, Mhatre-Winters I, Richardson JR. Microglia in Aging and Alzheimer’s Disease: A Comparative Species Review. Cells 2021;10(5):1138. doi: 10.3390/cells10051138. PMID: 34066847; PMCID: PMC8150617.

3. Ganguly U, Kaur U, Chakrabarti SS, et al. Oxidative Stress, Neuroinflammation, and NADPH Oxidase: Implications in the Pathogenesis and Treatment of Alzheimer’s Disease. Oxid Med Cell Longev 2021;2021:7086512. doi: 10.1155/2021/7086512. PMID: 33953837; PMCID: PMC8068554.

4. Price BR, Johnson LA, Norris CM. Reactive astrocytes: The nexus of pathological and clinical hallmarks of Alzheimer’s disease. Ageing Res Rev 2021;68:101335. doi: 10.1016/j.arr.2021.101335. Epub 2021 Mar 31. PMID: 33812051; PMCID: PMC8168445.

5. Silagi ML, Bertolucci PH, Ortiz KZ. Naming ability in patients with mild to moderate Alzheimer’s disease: what changes occur with the evolution of the disease? Clinics (Sao Paulo) 2015;70(6):423–8. doi: 10.6061/clinics/2015(06)07. Epub 2015 Jun 1. PMID: 26106961; PMCID: PMC4462568.

6. Aniwattanapong D, Tangwongchai S, Supasitthumrong T, et al. Validation of the Thai version of the short Boston Naming Test (T-BNT) in patients with Alzheimer’s dementia and mild cognitive impairment: clinical and biomarker correlates. Aging Ment Health 2019;23(7):840–50. doi: 10.1080/13607863.2018.1501668. Epub 2018 Oct 23. PMID: 30351202.

7. Tangwongchai S, Supasitthumrong T, Hemrunroj S, et al. In Thai Nationals, the ApoE4 Allele Affects Multiple Domains of Neuropsychological, Biobehavioral, and Social Functioning Thereby Contributing to Alzheimer’s Disorder, while the ApoE3 Allele Protects Against Neuropsychiatric Symptoms and Psychosocial Deficits. Mol Neurobiol 2018;55(8):6449–62. doi: 10.1007/s12035-017-0848-0. Epub 2018 Jan 6. PMID: 29307083.

8. Bettcher BM, Olson KE, Carlson NE, et al. Astrogliosis and episodic memory in late life: higher GFAP is related to worse memory and white matter microstructure in healthy aging and Alzheimer’s disease. Neurobiol Aging 2021;103:68–77. doi: 10.1016/j.neurobiolaging.2021.02.012. Epub 2021 Feb 26. PMID: 33845398.

9. Morris G, Berk M, Maes M, et al. Could Alzheimer’s Disease Originate in the Periphery and If So How So? Mol Neurobiol 2019;56(1):406–34. doi: 10.1007/s12035-018-1092-y. Epub 2018 Apr 29. PMID: 29705945; PMCID: PMC6372984.

10. Liu CC, Liu CC, Kanekiyo T, et al. Apolipoprotein E and Alzheimer disease: risk, mechanisms and therapy. Nat Rev Neurol 2013;9(2):106–18. doi: 10.1038/nrneurol.2012.263.

11. Butterfield DA, Mattson MP. Apolipoprotein E and oxidative stress in brain with relevance to Alzheimer’s disease. Neurobiol Dis 2020;138:104795. doi: 10.1016/j.nbd.2020.104795. Epub 2020 Feb 6. PMID: 32036033; PMCID: PMC7085980.

12. Kloske CM, Wilcock DM. The Important Interface Between Apolipoprotein E and Neuroinflammation in Alzheimer’s Disease. Front Immunol 2020;11:754. doi: 10.3389/fimmu.2020.00754. PMID: 32425941; PMCID: PMC7203730.

13. Supasitthumrong T, Tunvirachaisakul C, Aniwattanapong D, et al. Peripheral Blood Biomarkers Coupled with the Apolipoprotein E4 Genotype Are Strongly Associated with Semantic and Episodic Memory Impairments in Elderly Subjects with Amnestic Mild Cognitive Impairment and Alzheimer’s Disease. J Alzheimers Dis 2019;71(3):797–811. doi: 10.3233/JAD-190114. PMID: 31424390.

14. Lennon MJ, Makkar SR, Crawford JD, et al. Midlife Hypertension and Alzheimer’s Disease: A Systematic Review and Meta-Analysis. J Alzheimers Dis 2019;71(1):307–16. doi: 10.3233/JAD-190474. PMID: 31381518.

15. Lee SH, Han K, Cho H, et al. Variability in metabolic parameters and risk of dementia: a nationwide population-based study. Alzheimers Res Ther 2018;10(1):110. doi: 10.1186/s13195-018-0442-3. PMID: 30368247; PMCID: PMC6204276.

16. Carranza-Naval MJ, Vargas-Soria M, Hierro-Bujalance C, et al. Alzheimer’s Disease and Diabetes: Role of Diet, Microbiota and Inflammation in Preclinical Models. Biomolecules 2021;11(2):262. doi: 10.3390/biom11020262. PMID: 33578998; PMCID: PMC7916805.

17. Lee HJ, Seo HI, Cha HY, et al. Diabetes and Alzheimer’s Disease: Mechanisms and Nutritional Aspects. Clin Nutr Res 2018;7(4):229–40. doi: 10.7762/cnr.2018.7.4.229. Epub 2018 Oct 23. PMID: 30406052; PMCID: PMC6209735.

18. Petersen RC. Mild Cognitive Impairment. Continuum (Minneap Minn) 2016;22(2 Dementia):404–18. doi: 10.1212/CON.0000000000000313. PMID: 27042901; PMCID: PMC5390929.

19. Brambati SM, Belleville S, Kergoat MJ, et al. Single-and multiple-domain amnestic mild cognitive impairment: two sides of the same coin? Dement Geriatr Cogn Disord 2009;28(6):541–9. doi: 10.1159/000255240. Epub 2009 Dec 14. PMID: 20016185.

20. Michaud TL, Su D, Siahpush M, et al. The Risk of Incident Mild Cognitive Impairment and Progression to Dementia Considering Mild Cognitive Impairment Subtypes. Dement Geriatr Cogn Dis Extra 2017;7(1):15–29. doi: 10.1159/000452486. PMID: 28413413; PMCID: PMC5346939.

21. Simeonova D, Stoyanov D, Leunis JC, et al. Construction of a nitro-oxidative stress-driven, mechanistic model of mood disorders: A nomothetic network approach. Nitric Oxide 2021;106:45–54. doi: 10.1016/j.niox.2020.11.001. Epub 2020 Nov 10. PMID: 33186727.

22. Maes M, Moraes JB, Bonifacio KL, et al. Towards a new model and classification of mood disorders based on risk resilience, neuro-affective toxicity, staging, and phenome features using the nomothetic network psychiatry approach. Metab Brain Dis 2021;36(3):509–21. doi: 10.1007/s11011-020-00656-6. Epub 2021 Jan 7. PMID: 33411213.

23. Stoyanov D, Maes MH. How to construct neuroscience-informed psychiatric classification? Towards nomothetic networks psychiatry. World J Psychiatry 2021;11(1):1–12. doi: 10.5498/wjp.v11.i1.1. PMID: 33511042; PMCID: PMC7805251.

24. Morris JC. The Clinical Dementia Rating (CDR): current version and scoring rules. Neurology 1993;43(11):2412–4. doi: 10.1212/wnl.43.11.2412-a. PMID: 8232972.

25. Medical technology assessment project committee, The Comparison of the Test Performance Between the MMSE-Thai 2002 and the TMSE for Dementia Screening in the Elderly. 2008, Bangkok, Thailand: Thai Geriatric Medicine Institute, Ministry of Public Health.

26. Folstein MF, Folstein SE, McHugh PR. “Mini-mental state”. A practical method for grading the cognitive state of patients for the clinician. J Psychiatr Res 1975;12(3):189–98. doi: 10.1016/0022-3956(75)90026-6. PMID: 1202204.

27. McKhann G, Drachman D, Folstein M, et al. Clinical diagnosis of Alzheimer’s disease: report of the NINCDS-ADRDA Work Group under the auspices of Department of Health and Human Services Task Force on Alzheimer’s Disease. Neurology 1984;34(7):939–44. doi: 10.1212/wnl.34.7.939. PMID: 6610841.

28. Crowe M, Andel R, Wadley V, et al. Subjective cognitive function and decline among older adults with psychometrically defined am-nestic MCI. Int J Geriatr Psychiatry 2006; 21:1187–1192.

29. Morris JC, Heyman A, Mohs RC, et al. The Consortium to Establish a Registry for Alzheimer’s Disease (CERAD). Part I. Clinical and neuropsychological assessment of Alzheimer’s disease. Neurology 1989;39(9):1159–65. doi: 10.1212/wnl.39.9.1159. PMID: 2771064.

30. Benjamini Y, Hochberg Y. Controlling the false discovery rate: a practical and powerful approach to multiple testing. J Royal Statistics Soc Series b (Methodological) 1995;57:289–300.

31. Ringle CM, da Silva D, Bido, D. Structural equation modeling with the SmartPLS. Brazilian Journal of Marketing - BJM Revista Brasileira de Marketing – ReMark Edição Especial 2014, 13, n2.

32. Tunvirachaisakul C, Supasitthumrong T, Tangwongchai S, et al. Characteristics of Mild Cognitive Impairment Using the Thai Version of the Consortium to Establish a Registry for Alzheimer’s Disease Tests: A Multivariate and Machine Learning Study. Dement Geriatr Cogn Disord 2018;45(1-2):38–48. doi: 10.1159/000487232. Epub 2018 Apr 4. PMID: 29617684.

33. Hemrungrojn S, Tangwongchai S, Charoenboon T, et al. Use of the Montreal Cognitive Assessment Thai version (MoCA) to discriminate amnestic mild cognitive impairment from Alzheimer’s disease and healthy controls: machine learning results. Running head: MoCA and amnestic mild cognitive impairment. Preprints, May 2021.

34. Ismail Z, Smith EE, Geda Y, et al. Neuropsychiatric Symptoms Professional Interest Area. Neuropsychiatric symptoms as early manifestations of emergent dementia: Provisional diagnostic criteria for mild behavioral impairment. Alzheimers Dement 2016;12(2):195–202. doi: 10.1016/j.jalz.2015.05.017. Epub 2015 Jun 18. PMID: 26096665; PMCID: PMC4684483.

35. Johansson M, Stomrud E, Insel PS, et al. Mild behavioral impairment and its relation to tau pathology in preclinical Alzheimer’s disease. Transl Psychiatry 2021;11(1):76. doi: 10.1038/s41398-021-01206-z. PMID: 33500386; PMCID: PMC7838407.

